# CD19 CAR T-cell (BY19) therapy for Pediatric and Adult Patients with Relapsed or Refractory B-Cell Neoplasms in Belarus: Phase 1 trial

**DOI:** 10.1101/2025.08.20.25333782

**Authors:** Mikalai Katsin, Dmitri Dormeshkin, Alexandr Migas, Olga Karas, Tatsiana Shman, Yuliya Serada, Yauheniya Khalankova, Hanna Klych, Dzmitry Lutskovich, Alena Lukoika, Alexander Meleshko

## Abstract

Despite the approval of multiple CAR T-cell products, access to this therapy remains limited in many developing countries. We conducted a single-arm, open-label, non-randomized, parallel phase 1/2 clinical trial (ClinicalTrials.gov identifier: NCT05333302) at two independent centers: the Vitebsk Regional Clinical Cancer Centre (VRCCC) and the Belarusian Research Center for Pediatric Oncology, Hematology, and Immunology (BRC POHI). The study enrolled patients with relapsed or refractory B-cell malignancies who received an in-house manufactured CD19 CAR T-cell product (BY19) following lymphodepletion with either fludarabine plus cyclophosphamide or cyclophosphamide plus fludarabine and decitabine. Seven patients at VRCCC and sixteen at BRC POHI received CD19 CAR T-cell therapy, comprising 17 patients with B-ALL, one with chronic lymphocytic leukemia (CLL), and five with non-Hodgkin lymphoma. The median age was 45 years (range: 38–56) at VRCCC and 13.5 years (range: 4–30) at BRC POHI. Cytokine release syndrome (CRS) occurred in 18 (67%) of the 27 infusions across both centers, predominantly mild cases. Immune effector cell-associated neurotoxicity syndrome (ICANS) was observed in 12 (44%) patients, with severe ICANS (grade ≥3) in 5 patients (18.5%). The overall response rate (ORR) was 80.0% (16/20), with a complete response (CR) rate of 75.0% (15/20) at the first assessment on day 28. The median progression-free survival (PFS) was 23 months, and the median overall survival (OS) was 55 months. At 12 months, PFS was 83.3% for non-Hodgkin lymphoma patients and 48.3% for B-ALL patients. Higher Cmax levels tended to correlate with better response rates; however, no clear advantage in PFS was observed. In conclusion, our in-house manufactured CD19 CAR T-cell product (BY19) demonstrates a safety and efficacy profile comparable to approved CD19 CAR T-cell therapies. This study underscores the translational potential of localized CAR T-cell manufacturing to expand global access to advanced immunotherapies, especially in middle-income countries. Additionally, incorporating decitabine into the lymphodepletion regimen shows promise in enhancing therapeutic efficacy and warrants further prospective investigation.

## INTRODUCTION

Approximately 10% of children and young adults treated for upfront B-cell acute lymphoblastic leukemia (B-ALL) relapses or become refractory. The five-year overall survival (OS) rate following first relapse in this population is approximately 50% [1,2]. This rate drops to below 40% when patients have measurable residual disease (MRD)-positive status at the end of consolidation therapy [3]. In adults, the prognosis is considerably worse, with fewer than 10% of patients achieving long-term survival [4].

Diffuse large B-cell lymphoma (DLBCL) is the most common subtype of non-Hodgkin lymphoma. While first-line treatment yields five-year survival rates between 60% and 70%, up to 50% of patients either become refractory or relapse after initial therapy. For refractory DLBCL, the objective response rate (ORR) to subsequent treatments is approximately 26%, with only about 7% achieving complete response (CR), and the median OS remains around 6.3 months [5]. There has been a pressing need for novel therapeutic approaches to improve outcomes in these patient populations.

The field of hematology has recently been revolutionized by the advent of chimeric antigen receptor T cells (CAR-T cells) for the treatment of hematological malignancies, and it continues to evolve rapidly. The earliest efforts date back to 1989 when Gross et al. fused the alpha or beta chains of T cell receptor (TCR) constant (C) domains with the heavy (VH) or light (VL) chains of antibody variable (V) domains. These novel chimeric receptors were capable of transmitting activation signals upon binding to surface antigens in an MHC-independent manner [6]. In 1993, Eshhar et al. developed the first-generation CAR by linking the single-chain variable fragment (scFv) of an antibody to intracellular signaling domains derived from either the CD3γ or CD3ζ chain [7]. Building upon this foundation, various research groups demonstrated that second-generation CAR-T cells offered enhanced functionality by incorporating an additional co-stimulatory domain—typically either CD28 or 4-1BB—alongside CD3ζ, leading to improved T cell activation and persistence [8, 9]. Currently, second-generation CD19 CAR-T cells remain the most extensively studied both preclinically and clinically for the treatment of hematological malignancies. CD19, a member of the immunoglobulin superfamily, plays a role in B cell receptor signaling and is expressed on the surface of B cells at most stages of their development [10]. It is considered an ideal target for CAR-T cell therapies because it is expressed on the majority of B cell malignancies while being absent in hematopoietic stem cells. B-cell aplasia is a manageable on-target treatment effect associated with CD19-directed CAR-T therapy [11]. However, a potential limitation of targeting CD19 is that it is not involved in maintaining the tumorigenic phenotype, and escape variants—such as CD19-negative relapses have been reported [12].

Tisagenlecleucel, an autologous first-in-class anti-CD19 CAR-T cell product, has been evaluated for children and young adults (defined as those aged ≤25 years) with relapsed or refractory (R/R) B-ALL in the global phase II ELIANA clinical trial. Long-term follow-up of these patients demonstrated an overall CR rate of 82% and a median event-free survival (EFS) of 24 months. The estimated 3-year relapse-free survival rates were 52% when censoring for subsequent therapies and 48% without such censoring. These promising results led to the U.S. Food and Drug Administration (FDA) approval of Tisagenlecleucel, the first CAR-T cell therapy approved for children and young adults with R/R B-ALL, which is marketed by Novartis [13]. Other studies have reported similar outcomes, with MRD-negative CR achieved in 60% to 93% of cases [14–17].

For DLBCL, CR rates of 40-50% have been reported, whereas in patients with chronic lymphocytic leukemia (CLL), CR rates of 15-30% have been observed [18]. To date, five CD19 CAR-T cell products have received approval from the FDA for various B-cell malignancies, highlighting the significant success and therapeutic potential of this modality.

However, widespread clinical adoption is limited by high manufacturing costs, centralized production logistics, lengthy turnaround times, and complex regulatory requirements—factors that collectively restrict access, particularly in low- and middle-income countries.

To address this critical gap, we have developed an anti-CD19 CAR design that incorporates an IgG4 hinge and a 4-1BB cytoplasmic domain for our clinical program targeting patients with B-cell neoplasms. Considering the immunological benefits of decitabine observed in enhancing CD19 CAR T-cell activity against B-ALL and non-Hodgkin lymphomas, we have included decitabine in the lymphodepletion regimen for adult patients with B-cell neoplasms undergoing CD19 CAR T-cell therapy [19, 20].

Here, we present the results of a phase I study (ClinicalTrials.gov identifier: NCT05333302) evaluating CD19 CAR-T cells (BY19) in patients with relapsed or refractory B-cell malignancies conducted in the Republic of Belarus, demonstrating both the feasibility of manufacturing and preliminary durable responses across different B-cell neoplasms.

This study represents the first formal clinical investigation of an academic CAR T-cell product in Eastern Europe and provides important translational insight into expanding access to CAR T-cell therapies in resource-constrained healthcare systems.

## METHODS

### CAR expression cassettes

The BY19 CAR cassette encodes a single-chain variable fragment (scFv) derived from the CD19 antibody (FMC63), preceded by the CSF2R signal peptide and linked to the short hinge (12 amino acids) derived from human IgG4 domain. This is followed by the transmembrane domain of CD28, the 4-1BB co-stimulatory domain, and the CD3ζ (zeta) signaling chain.

For comparison, a CD28-based CD19 CAR (28Z) was designed to mimic the Axicabtagene ciloleucel configuration. This construct contains the FMC63 scFv linked to the CD28 hinge, transmembrane, and co-stimulatory domains, followed by the CD3ζ signaling chain. Both CAR constructs include a membrane-anchored truncated EGFR domain separated from the CD3ζ chain by a GSG linker and a P2A self-cleaving peptide sequence.

### Production of Recombinant Lentiviral Particles

Recombinant lentiviral particles were produced by transient transfection of 293T packaging cells with the second-generation transfer vector pWPXL (encoding the respective expression cassettes), the packaging plasmid pCMV-dR8.91, and the envelope plasmid pMD2.G, using linear polyethylenimine (Serochem, Prime-AQ100-100ML). 48 h post-transfection virus-containing supernatants were collected and clarified by centrifugation, filtered through 0.45 µm syringe filters, and concentrated by low-speed centrifugation (3000 × g, 4 °C, 24 h). Viral pellets were resuspended in ImmunoCult™-XF T Cell Expansion Medium (STEMCELL Technologies, Inc., 10981) and stored at −80 °C. Functional titers were determined by transducing 293T cells with serial dilutions of the viral preparation, followed by flow cytometric analysis of transduction efficiency.

### Cell lines and general cell culture

B-cell malignant cell lines Raji (ATCC CCL-86) and CII (DSMZ ACC-773), chronic myelogenous leukemia cell line K562 (ATCC CCL-242), embryonic kidney cell line 293T (ATCC CRL-3216) and T-cell leukemia cell line Jurkat E6.1 (ATCC TIB-152) were originally purchased from the American Type Culture Collection (ATCC) or the Deutsche Sammlung von Mikroorganismen und Zellkulturen (DSMZ). The reporter cell lines Jurkat_NFAT.GFP and Jurkat_NFkB.GFP, derivatives of the Jurkat E6.1 line, carry reporter constructs that enable expression of green fluorescent protein (GFP) in response to activation of the respective signaling pathways. These lines were generated in-house as previously described [21]. The cell lines K562, Raji, and CII were stably transduced to express the red fluorescent protein iRFP713, which served as a unique identifier for flow cytometric analysis in mixed cultures. All cell lines were maintained in accordance with the respective collection guidelines.

### Generation of CAR T-cells for Clinical Use

Peripheral blood mononuclear cells (PBMCs) were isolated from leukapheresis products of enrolled patients by density gradient centrifugation using ROTI®-Sep 1077 human solution (Carl Roth GmbH + Co. KG, 0642.2). In seven cases, the PBMC fraction was cryopreserved; in the remaining cases, cells were processed immediately for CAR T-cells manufacture. CD4□ and CD8□ T-lymphocytes were manually isolated using Dynabeads CD4 and CD8 Positive Isolation Kits (ThermoFisher Scientific, 11331D and 11333D). Two patients (Pt03, Pt05) underwent immunomagnetic selection of naive T cells EasySep™ Human Naïve CD4/8+ T Cell Isolation Kits (STEMCELL Technologies, Inc., 19555 and 19258) for CAR-T production.

Following enrichment, CD4□ and CD8□ T-cells were separately stimulated with CTS Dynabeads CD3/CD28 (ThermoFisher Scientific, 40203D). 48h post-activation, T-cells were genetically modified with pre-generated recombinant lentiviral particles encoding BY19 expression cassette via spinoculation on RetroNectin (Takara Bio, Inc., T202) - coated plates at a multiplicity of infection (MOI) of 5. Activation, transduction, and subsequent expansion were performed in ImmunoCult™-XF T Cell Expansion Medium (STEMCELL Technologies, Inc., 10981) supplemented with recombinant human interleukin-7 (Miltenyi Biotec, 170-076-111) and interleukin-15 (Miltenyi Biotec, 170-076-114) each at a final concentration of 10 ng/mL.

### Generation of CAR T-cells and CAR-Expressing Jurkat Reporter Lines for in vitro tests

Primary CD4□ and CD8□ T-lymphocytes were isolated from the peripheral blood of a healthy donor by immunomagnetic selection (Miltenyi Biotec, 130-096-535; 130-042-201). T-cells were activated using magnetic beads (Miltenyi Biotec, 130-091-441). 48 h post-activation, T-cells were transduced with pre-generated recombinant lentiviral particles encoding BY19 or 28Z expression cassettes via spinoculation in the presence of Vectofusin-1 transduction enhancer (Miltenyi Biotec, 130-111-163) at MOI of 5. Cells were expanded for 12–14 days in RPMI-1640 medium (Capricorn, RPMI-HA) supplemented with 10% FBS (Capricorn, FBS-HI-12A), recombinant human interleukin-7 (Miltenyi Biotec, 130-095-362), and interleukin-15 (Miltenyi Biotec, 130-095-764), each at a final concentration of 10 ng/mL.

Genetic modification of the parental Jurkat E6.1 cell line and its reporter derivatives, Jurkat_NFAT.GFP and Jurkat_NFkB.GFP, was performed using pre-generated recombinant lentiviral particles encoding BY19 or 28Z expression cassettes at MOI of 0.2.

### Flow cytometry

Flow cytometry acquisition was performed with a DxFlex flow cytometer (Beckman Coulter, C78505). Data analysis was performed using CytExpert software (Beckman Coulter, CytExpert for DxFLEX 2.0).

CARs identification on the surface of CAR T-cells was performed by biotinylated recombinant CD19 protein (in-house production) and streptavidin-APC conjugate (BioLegend, 405207). tEGFR reporter was identified by AlexaFluor 488-conjugated biosimilar of cetuximab (RnD Systems, FAB9577G-100).

The phenotypic composition and quantity of T-lymphocytes in the patient’s blood before apheresis, PBMCs and final CAR-T product were determined by staining the cells with antibodies CD45 (Beckman Coulter. c.J33), CD4 (Invitrogen, c.RPA-T4), CD8 (Invitrogen, c.RPA-T8), CD3 (Beckman Coulter, c.UCHTI), CD45RO (Invitrogen, c.HI100), CCR7 (Miltenyi Biotec, c.REA546). CAR-T cell persistence after infusion in blood, bone marrow and cerebrospinal fluid was monitored using antibodies to CD45 (Beckman Coulter. c.J33), CD4 (Invitrogen, c.RPA-T4), CD8 (Invitrogen, c.RPA-T8), CD3 (Beckman Coulter, c.UCHTI), and human EGFRt cetuximab biosimilar (Invitrogen c.Me183). DAPI (Invitrogen, 62248) was used as a viability dye.

### In vitro assessment of CAR Functionality

To assess CAR surface expression, Jurkat E6.1 cells were transduced with recombinant lentiviral particles carrying the corresponding expression cassettes. Genetically modified cells were stained with recombinant CD19 protein to detect the FMC63-based scFv and with a cetuximab biosimilar to detect the tEGFR reporter. CAR expression was normalized by calculating the ratio of the FMC63 scFv mean fluorescence intensity (MFI) to that of tEGFR.

To evaluate the functional activity of the CAR intracellular signaling domains, reporter cell lines Jurkat_NFAT.GFP and Jurkat_NFkB.GFP expressing the analyzed CARs were co-cultured with target cells (K562, Raji, CII) at an effector-to-target (E:T) ratio of 1:1 for 24 h. GFP reporter expression was measured by flow cytometry at 0, 4, and 24 h.

### In vitro assessment of Primary CAR T-cells Functional Activity

Primary CAR T cells, generated from healthy donor material, were incubated for 48 h in complete medium without adding cytokines prior to assays initiation. Cells were then washed and mixed with non-transduced T cells to adjust the final CAR-positive fraction to 25%.

For the 24 h co-cultivation assay, effector cells were mixed with target cells (K562, Raji, CII) at effector-to-target (E:T) ratios of 1:1, and incubated for 24 h prior to flow cytometric analysis. The fold expansion of target cells was determined by dividing the number of live target cells detected after 24 h of co-cultivation with CAR T cells by the number of input target cells at assay initiation.

For the rechallenge assay, effector cells were co-cultured with target cells (Raji, CII) at an E:T ratio of 1:1. Every 48–72 h, cell content was analyzed by flow cytometry and fresh live target cells were added in the same quantity as the initial input. A total of five restimulation rounds were performed.

Target cell content was determined by flow cytometry based on iRFP713 expression, and CAR T cell content was determined based on tEGFR expression.

CAR-T culture supernatants were assessed for IFNγ production after 24 h of co-cultivation with target cells using an ELISA kit (ElabScience, E-UNEL-H0069), following the manufacturer’s instructions.

### Study design and participants

Adult and pediatric patients were enrolled in a single-arm, an open-label, non-randomized, parallel phase I/II clinical trial (ClinicalTrials.gov identifiers: NCT05333302) conducted at two independent centers: Vitebsk Regional Clinical Cancer Centre (VRCCC) and the Belarusian Research Center for Pediatric Oncology, Hematology and Immunology (BRC POHI). The study was approved by the respective institutional review boards and conducted in accordance with the Declaration of Helsinki and International Conference on Harmonization guidelines for Good Clinical Practice. All patients provided written informed consent. Patients with R/R CD19+ lymphoid neoplasms received autologous or allogeneic anti-CD19 CAR-T cells manufactured at BRC POHI, utilizing clinical-grade lentiviral vectors and following standard operating procedures for cell production. At VRCCC, CAR T-cell products - either fresh or thawed, were administered within 4 hours of preparation. Eligible patients were required to have a life expectancy of at least 8 weeks, have recovered from acute toxic effects of prior chemotherapy, immunotherapy, or radiotherapy, and demonstrate adequate organ function. An absolute T-cell count of ≥150 cells/µL was also required. Patients who had undergone allogeneic hematopoietic stem cell transplantation (alloSCT) needed to have confirmed CD19+ leukemia recurrence (defined as ≥0.01% disease by multiparameter flow cytometry), be free from active graft-versus-host disease (GVHD) and have discontinued immunosuppressive therapy for at least 4 weeks prior to enrollment. Patients with extramedullary disease or central nervous system (CNS) leukemic involvement were eligible if asymptomatic.

Bridging chemotherapy was permitted prior to lymphodepletion. For patients enrolled at VRCCC, the lymphodepletion regimen consisted of cyclophosphamide at 250 mg/m² and fludarabine at 25 mg/m² administered on days −5 to −3, along with decitabine at the full dose of 100 mg/m² on days −6 to −4. A BRC POHI, the lymphodepletion included full doses of cyclophosphamide at 750 mg/m² and fludarabine at 120 mg/m² given on days −4 to −2. The target dose of CD19 CAR-T cells for patients at VRCCC was 50–70 × 10^6 cells. Conversely, at BRC POHI, the administered dose ranged from a minimum of 0.1 × 10^6 CAR-positive T-cells per kilogram of body weight (up to a maximum of 2.76 × 10^6 cells/kg). Tocilizumab was administered prior to infusion to manage cytokine release syndrome. Additionally, a second infusion of CD19 CAR-T cells was permitted for patients at BRC POHI (Figure 1).

**Figure 1.**
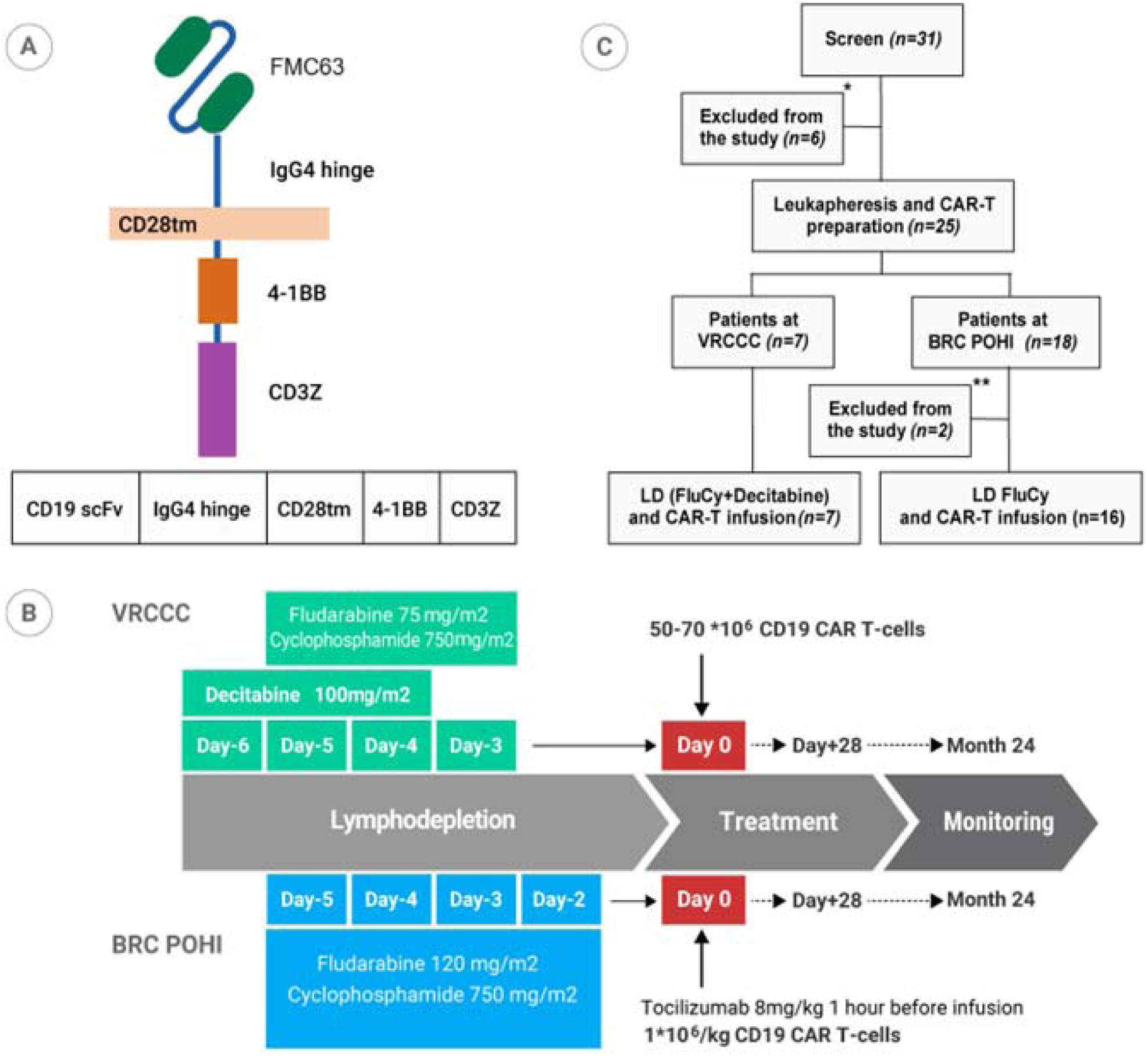
A. Anti-CD19 CAR design B. Design of the phase 1/2 clinical trial NCT05333302 C. Flowchart for CD19 CAR-T cells clinical trial including cell procurement and treatment. Seven patients received lymphodepletion with fludarabine, cyclophosphamide and decitabine with subsequent anti-CD19 CAR-T cells infusion at Vitebsk regional clinical Cancer Centre (VRCCC). Sixteen patients received standard fludarabine, cyclophosphamide lymphodepletion and infusion of anti-CD19 CAR-T cells at Belarusian Research Center for Pediatric Oncology, Hematology and Immunology (BRC POHI).

The primary endpoint was safety. Secondary endpoints included ORR, CR, progression-free survival (PFS), OS, as well as the expansion and persistence of CAR-T cells. Cytokine release syndrome (CRS) and immune effector cell-associated neurotoxicity syndrome (ICANS) were graded according to the consensus criteria of the American Society for Transplantation and Cellular Therapy (ASTCT) [22). Hemophagocytic lymphohistiocytosis (HLH) was graded based on Shah N.N. criteria [23]. Immune effector cell-associated hematotoxicity (ICAHT) grading followed the EHA/EBMT consensus guidelines [24]. All other toxicities were graded using the National Cancer Institute’s Common Terminology Criteria for Adverse Events, version 4.0.

Response assessment for lymphoma and B-ALL with extramedullary involvement was performed according to the Lugano classification 2014, utilizing FDG PET-CT or CT scans at +28 days post-treatment and subsequently every 3 months [25]. Brain MRI was employed to evaluate CNS involvement. Responses in B-ALL were graded according to standard criteria, while CLL/SLL (small lymphocytic lymphoma) responses were assessed based on the iwCLL 2018 guidelines [26].

### Statistical analysis

All in vitro assays were performed in triplicates. Data are presented as mean ± standard deviation (SD). Group comparisons were performed using ordinary one-way ANOVA, with differences considered statistically significant at a p-value <0.05. Statistical analyses and data visualization were conducted using GraphPad Prism software, version 8 (GraphPad Software, San Diego, CA, USA).

Standard descriptive statistics, including median/range, percentage, and response rate, were reported for key variables for clinical results assessment. Directional associations between categorical variables with three levels and various predictors were evaluated using univariate proportional odds logistic regression. A p-value <0.05 was considered statistically significant.

Kaplan-Meier curves were generated for overall survival (OS), progression-free survival (PFS). The OS and PFS curves included data from all infused patients. Log-rank tests were performed to assess significance between these groups.

For the engraftment analysis, peak engraftment (expressed as a percentage or absolute number of total T cells) and time-averaged engraftment (measured as the area under the curve) from infusion to day +90 were calculated for each patient. These values were then grouped by risk factors as outlined in this section, and differences were evaluated using a two-sided Wilcoxon-Mann-Whitney test (for comparisons between two groups) or a Kruskal-Wallis test (for comparisons involving three or more groups).

## RESULTS

### Despite exhibiting lower CAR surface expression, BY19 CD19 CAR T-cells demonstrate greater proliferation and tumor control compared to CD28-based CAR T-cells in preclinical models

We constructed two of the most widely used designs of second-generation anti-CD19 CARs: BY19 CAR, which incorporates an IgG4 hinge, CD28 transmembrane domain, 4-1BB cytoplasmic domain, and CD3ζ signaling domain; and a second 28Z CAR possessing CD28 hinge, transmembrane, and cytoplasmic domains. The surface expression of CARs on T-cells was assessed by staining with recombinant CD19 protein, followed by flow cytometric analysis. The tEGFR reporter, included in the expression cassettes, was detected using a cetuximab biosimilar (Figure 2A). BY19 CAR T-cells demonstrated lower surface expression compared to the CD28 comparator (Figure 2B). The signaling strength and activation of the BY19 and CD28Z designs were evaluated using NFAT and NF-κB reporter assays with Raji and CII (CD19+) cells as target cells, and K562 cells serving as a negative control. Despite exhibiting lower surface CAR expression, BY19 CAR T-cells demonstrated statistically significant enhanced activation signaling in both assays across both cell lines (Supplementary figure 1). To evaluate the cytotoxic activity of CD19 CAR T-cells, Raji and CII cancer cell lines were used (Figure 2C). In short-term cytotoxicity assays, both CAR T-cell designs (BY19 and CD28Z) exhibited similar levels of cytotoxicity, although CD28Z CAR T-cells tended to produce higher levels of IFN-γ (Figure 2 C, D).

**Figure 2.**
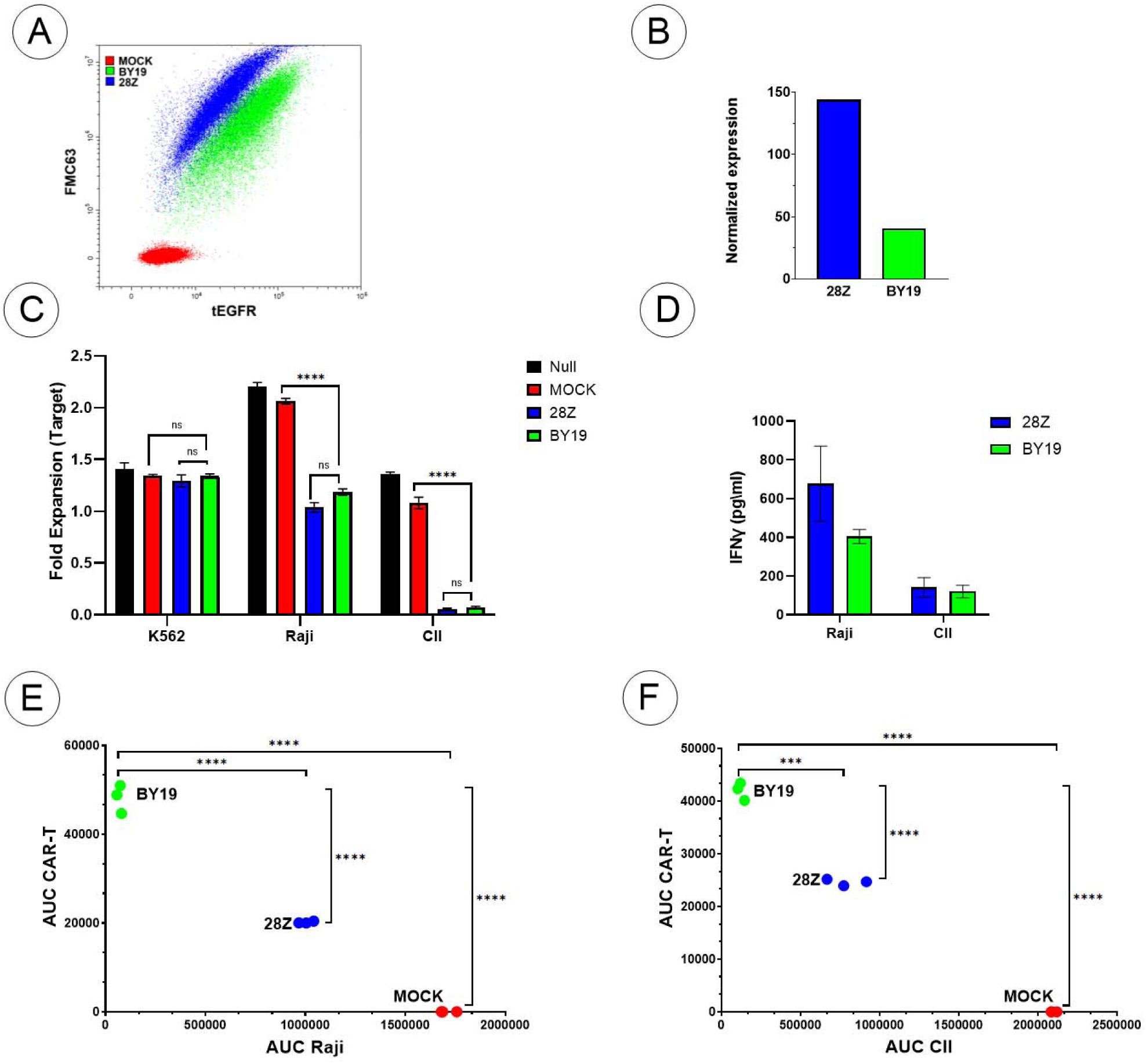
-In vitro characterization and cytotoxicity assessment of BY19 CAR T-cells compared to CD28Z CAR T-cells. A) CAR expression of transduced T-cells. Extracellular modules of BY19 and 28Z were directly stained with biotin-labeled CD19 protein and AlexaFluor 647-conjugated streptavidin, and tEGFR reporter was stained with AlexaFluor 488-conjugated cetuximab biosimilar. MOCK - untransduced cells; B) CAR expression normalized to tEGFR. BY19 and 28Z MFI levels were normalized to the MFI levels of tEGFR reporter protein expression; C) CAR mediated cytotoxic activity of T-cells in 24h co-cultivation assay. Fold expansion of target cells in short term co-cultivation assay with BY19 and 28Z CAR T-cells at E:T ratio 1:1. Data represent Mean ± SD. MOCK - co-culture of target cells with untransduced T-cells. Null - culture of target cells alone; D) ELISA-based quantification of IFN_γ_ production. Supernatants after 24h co-culture of engineered CAR T-cells with Raji or CII target cells. Data represent means ± SD; E) Control of Raji cells and proliferation of CAR T cells in rechallenge assay. AUC of target and CAR T cells in five rounds of rechallenge assay. MOCK - co-culture of target cells with untransduced T-cells; F) Control of CII cells and proliferation of CAR T cells in rechallenge assay. AUC of target and CAR T cells in five rounds of rechallenge assay. MOCK - co-culture of target cells with untransduced T-cells.

It is well established that the ability of CAR T-cells to maintain their cytotoxic capacity in the context of high tumor burden and repeated antigen stimulation correlates with clinical efficacy. To compare the functional persistence and cancer cell killing capacity of BY19 and CD28Z CAR T-cells, we performed a long-term rechallenge assay. After five rounds of re-exposure to CD19+ cancer cell lines (Raji and CII), BY19 CAR T-cells were superior in terms of specific cytolytic activity and proliferation (Figure 2 E, F). On the basis of preclinical evaluation, BY19 CAR T-cells were identified to have potent and antigen-specific anti-leukemia activity and were used in the subsequent clinical study.

### Patients Characteristics

Between September 2020 and May 2024, a total of 31 patients were screened, with 7 patients at VRCCC and 16 at BRC POHI ultimately receiving treatment between October 2020 and September 2024. Eight patients were not treated due to uncontrolled infectious complications and rapid disease progression; six of these patients died before peripheral blood could be collected for CAR T-cell manufacturing, and two died prior to CAR T-cell infusion. The median age was 45 years (range: 38–56) at VRCCC and 13.5 years (range: 4–30) at BRC POHI. At BRC POHI, 14 patients had B-ALL and two had Burkitt lymphoma. At VRCCC, one patient had CML in B-cell lymphoid blast crisis, two had B-ALL, one had primary mediastinal large B-cell lymphoma (PMBCL), one had DLBCL, one had primary CNS lymphoma (PCNSL), and one had CLL (see Table 1). The median number of prior therapy lines was 3 (range: 2–6). Seven out of sixteen patients (44%) with B-ALL underwent alloSCT, while eight patients (50%) received Blinatumomab prior to CD19 CAR-T cell infusion. Nineteen of twenty-three patients (83%) had a T-cell count exceeding 500 cells/μL. Extramedullary involvement was reported in 10 (55.5%) of 18 patients with B-ALL, with CNS involvement documented in one patient at the time of CD19 CAR T-cell infusion. Most B-ALL patients exhibited M1 bone marrow status (n=10), while M2 and M3 statuses were observed in four patients each. Bridge therapy between leukapheresis and lymphodepletion was utilized in seven patients—radiotherapy in two at VRCCC and chemotherapy in five at BRC POHI. Four patients with B-ALL received a second infusion of CD19 CAR-T cells at BRC POHI (see Figure 3).

**Figure 3.**
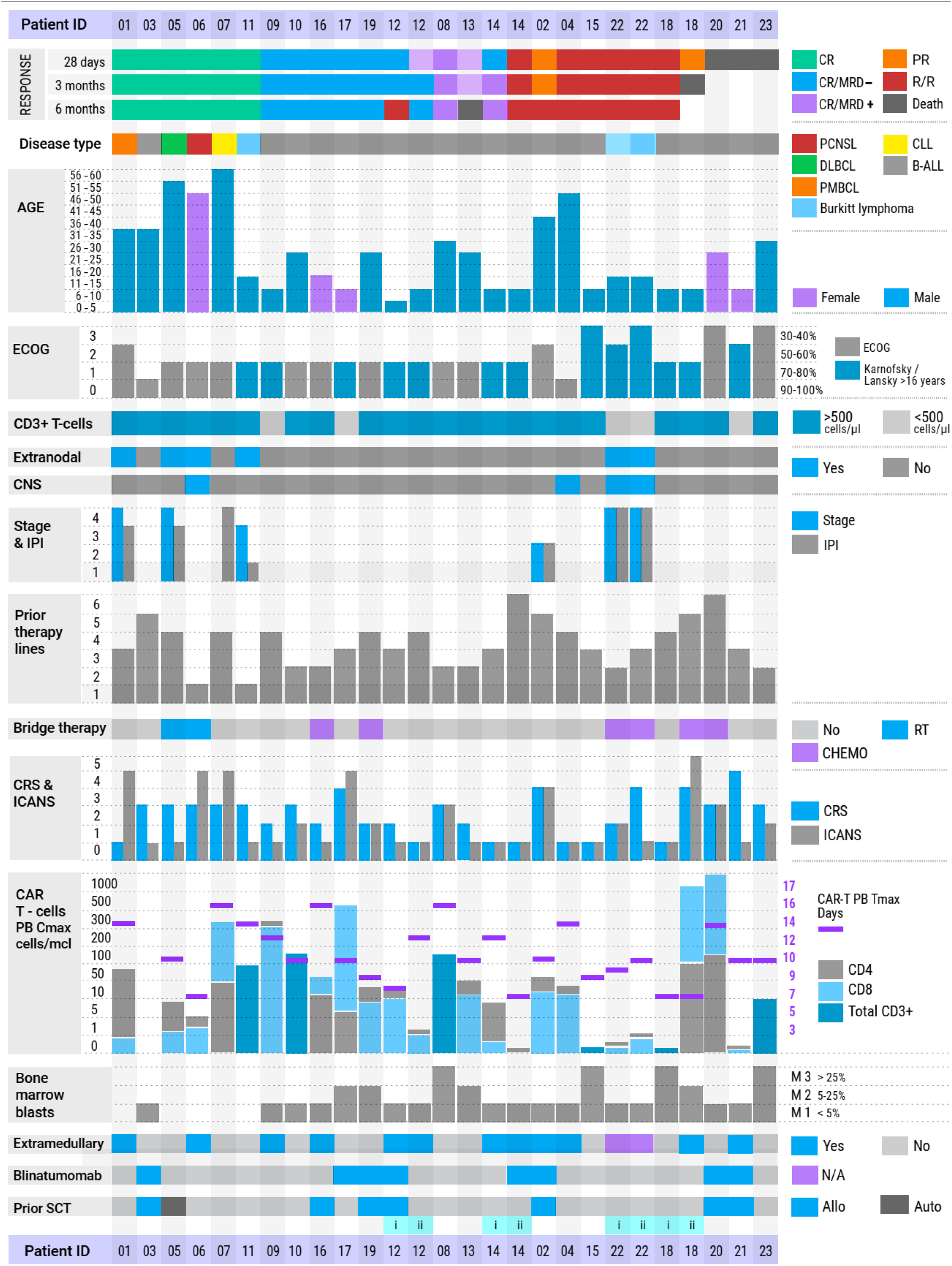
Summary of patients characteristics, toxicity and clinical response data (n=23) DLBCL, Diffuse large B-cell lymphoma; PCNSL, Primary CNS lymphoma; PMBCL, Primary mediastinal large B-cell lymphoma; CLL, chronic lymphocytic

**Table 1.** Adverse events occurred after anti-CD19 CAR-T cell therapy. VRCCC, Vitebsk regional clinical Cancer Centre; BRC POHI Belarusian Research Center for Pediatric Oncology, Hematology and Immunology; CRS, cytokine release syndrome; ICANS, Immune effector cell-associated neurotoxicity syndrome; HLH, hemophagocytic lymphohistiocytosis; ALT, Alanine Aminotransferase; AST, Aspartate Aminotransferase; ICAHT, Immune Effector Cell-Associated Hematotoxicity; LDH, Lactate Dehydrogenase; CRP, C-reactive protein; APTT, activated partial thromboplastin time.

| Treatment- related adverse events | VRCCC (n=7) |  |  |  | BRC POHI (n=20) |  |  |  |  | All infusions (n=27) |  |  |  |  |
| --- | --- | --- | --- | --- | --- | --- | --- | --- | --- | --- | --- | --- | --- | --- |
|  | Grade 1 | Grade 2 | Grade 3 | Grade 4 | Grade 1 | Grade 2 | Grade 3 | Grade 4 | Grade 5 | Grade 1 | Grade 2 | Grade 3 | Grade 4 | Grade 5 |
| CRS | 0 | 3 (42,8%) | 1 (14,3%) | 1 (14,3%) | 6 (30%) | 6 (30%) | 2 (10%) | 0 |  | 6 (22,2%) | 9 (33,3%) | 3 (11,1%) | 1 (3,7%) |  |
| ICANS | 0 | 0 | 0 | 3 (42,8%) | 6 (30%) | 2 (10%) | 0 | 1 (5%) | 1 (5%) | 6 (22,2%) | 2 (7,4%) | 0 | 4 (14,8%) | 1 (3,7%) |
| HLH | 0 | 0 | 0 | 2 (28,6%) | 0 | 0 | 0 | 0 | 1 (5%) | 0 | 0 | 0 | 2 (7,4%) | 1 (3,7%) |
| Infection | 2 (28,6%) | 0 | 1 (14,3%) | 1 (14,3%) | 0 | 0 | 3 (15%) | 1 (5%) | 2 (10%) | 2 (7,4%) | 0 | 4 (14,8%) | 2 (7,4%) | 2 (7,4%) |
| Chills | 2 (28,6%) | 0 | 0 | 0 | 1 (5%) | 4 (20%) | 0 | 0 |  | 3 (11,1%) | 4 (14,8%) | 0 | 0 |  |
| Pain |  |  |  |  | 2 (10%) | 5 (25%) | 2 (10%) | 0 |  | 2 (7,4%) | 5 (18,5%) | 2 (7,4%) | 0 |  |
| Fatigue | 3 (42,8%) | 0 | 2 (28,6%) | 0 | 6 (30%) | 9 (45%) | 0 | 0 |  | 9 (33,3%) | 9 (33,3%) | 2 (7,4%) | 0 |  |
| Hypotension | 0 | 1 (14,3%) | 3 (42,8%) | 1 (14,3%) | 0 | 4 (20%) | 2 (10%) | 4 (20%) |  | 0 | 5 (18,5%) | 5 (18,5%) | 5 (18,5%) |  |
| Tachycardia | 1 (14,3%) | 2 (28,6%) | 0 | 0 | 3 (15%) | 7 (35%) | 0 | 0 |  | 4 (14,8%) | 9 (33,3%) | 0 | 0 |  |
| Headache | 0 | 0 | 1 (14,3%) | 0 | 5 (25%) | 2 (10%) | 3 (15%) | 0 |  | 5 (18,5%) | 2 (7,4%) | 4 (14,8%) | 0 |  |
| ALT increased | 0 | 2 (28,6%) | 0 | 0 | 6 (30%) | 2 (10%) | 2 (10%) | 1 (5%) |  | 6 (22,2%) | 4 (14,8%) | 2 (7,4%) | 1 (3,7%) |  |
| AST increased | 1 (14,3%) | 1 (14,3%) | 1 (14,3%) | 0 | 8 (40%) | 3 (15%) | 2 (10%) | 2 (10%) |  | 9 (33,3%) | 4 (14,8%) | 3 (11,1%) | 2 (7,4%) |  |
| Anemia | 1 (14,3%) | 4 (57,1%) | 2 (28,6%) | 0 | 2 (10%) | 8 (40%) | 8 (40%) | 1 (5%) |  | 3 (11,1%) | 12 (44,4%) | 10 (37%) | 1 (3,7%) |  |
| Thrombocytopenia | 1 (14,3%) | 0 | 0 | 5 (71,4%) | 2 (10%) | 1 (5%) | 9 (45%) | 7 (35%) |  | 3 (11,1%) | 1 (3,7%) | 9 (33,3%) | 12 (44,4%) |  |
| ICAHT - Early | 0 | 0 | 4 (57,1%) | 3 (42,8%) |  |  |  | 19 (95%) |  | 0 | 0 | 4 (14,8%) | 22 (81,5%) |  |
| ICAHT - Late | 1 (14,3%) | 1 (14,3%) | 2 (28,6%) | 2 (28,6%) |  | 4 (20%) | 3 (15%) |  |  | 1 (3,7%) | 5 (18,5%) | 5 (18,5%) | 2 (7,4%) |  |
| Nausea | 0 | 7 (100%) | 0 | 0 | 9 (45%) | 1 (5%) | 0 | 0 |  | 9 (33,3%) | 8 (29,6%) | 0 | 0 |  |
| Vomiting | 1 (14,3%) | 3 (42,8%) | 0 | 0 | 5 (25%) | 2 (10%) | 0 | 0 |  | 6 (22,2%) | 5 (18,5%) | 0 | 0 |  |
| Diarrhea | 1 (14,3%) | 0 | 0 | 0 | 2 (10%) | 1 (5%) | 0 | 0 |  | 3 (11,1%) | 1 (3,7%) | 0 | 0 |  |
| Ataxia | 1 (14,3%) | 0 | 4 (57,1%) | 0 | 0 | 1 (5%) | 0 | 0 |  | 1 (3,7%) | 1 (3,7%) | 4 (14,8%) | 0 |  |
| Hypofibrinogenemia | 0 | 1 (14,3%) | 2 (28,6%) | 0 | 6 (30%) | 4 (20%) | 1 (5%) | 1 (5%) |  | 6 (22,2%) | 5 (18,5%) | 3 (11,1%) | 1 (3,7%) |  |
| Back pain |  |  |  |  | 0 | 3 (15%) | 0 | 0 |  | 0 | 3 (11,1%) | 0 | 0 |  |
| Cognitive disturbance | 0 | 0 | 3 (42,8%) | 0 | 0 | 1 (5%) | 0 | 0 |  | 0 | 1 (3,7%) | 3 (11,1%) | 0 |  |
| Confusion | 0 | 0 | 1 (14,3%) | 2 (28,6%) | 0 | 1 (5%) | 1 (5%) | 0 |  | 0 | 1 (3,7%) | 2 (7,4%) | 2 (7,4%) |  |
| Dizziness | 2 (28,6%) | 0 | 3 (42,8%) | 0 | 2 (10%) | 2 (10%) | 0 | 0 |  | 4 (14,8%) | 2 (7,4%) | 3 (11,1%) | 0 |  |
| Dysarthria | 1 (14,3%) | 0 | 2 (28,6%) | 0 | 0 | 1 (5%) | 0 | 0 |  | 1 (3,7%) | 1 (3,7%) | 2 (7,4%) | 0 |  |
| Dysphasia | 0 | 0 | 2 (28,6%) | 0 | 0 | 0 | 1 (5%) | 0 |  | 0 | 0 | 3 (11,1%) | 0 |  |
| Dyspnea | 2 (28,6%) | 1 (14,3%) | 0 | 0 | 1 (5%) | 4 (20%) | 2 (10%) | 0 |  | 3 (11,1%) | 5 (18,5%) | 2 (7,4%) | 0 |  |
| Fever | 2 (28,6%) | 3 (42,8%) | 0 | 0 | 2 (10%) | 2 (10%) | 0 | 0 |  | 4 (14,8%) | 5 (18,5%) | 0 | 0 |  |
| Elevated CRP | 0 | 2 (28,6%) | 0 | 5 (71,4%) | 10 (50%) | 0 | 0 | 0 |  | 10 (37%) | 2 (7,4%) | 0 | 5 (18,5%) |  |
| Elevated LDH | 4 (57,1%) | 0 | 0 | 0 | 12 (60%) | 0 | 0 | 0 |  | 16 (59,3%) | 0 | 0 | 0 |  |
| Hypoxia | 0 | 0 | 1 (14,3%) | 0 | 0 | 1 (5%) | 3 (15%) | 3 (15%) |  | 0 | 1 (3,7%) | 4 (14,8%) | 3 (11,1%) |  |
| Hypoalbuminemia | 2 (28,6%) | 0 | 0 | 0 | 5 (25%) | 1 (5%) | 0 | 0 |  | 7 (25,9%) | 1 (3,7%) | 0 | 0 |  |
| Hypokalemia | 2 (28,6%) | 2 (28,6%) | 1 (14,3%) | 0 | 0 | 0 | 5 (25%) | 0 |  | 2 (7,4%) | 2 (7,4%) | 6 (22,2%) | 0 |  |
| Hypocalcemia | 4 (57,1%) | 0 | 2 (28,6%) | 0 | 1 (5%) | 4 (20%) | 1 (5%) | 0 |  | 5 (18,5%) | 4 (14,8%) | 3 (11,1%) | 0 |  |
| APTT increased | 0 | 0 | 1 (14,3%) | 0 | 2 (10%) | 2 (10%) | 0 | 0 |  | 2 (7,4%) | 2 (7,4%) | 1 (3,7%) | 0 |  |
| Seizure |  |  |  |  | 0 | 0 | 1 (5%) | 1 (5%) |  | 0 | 0 | 1 (3,7%) | 1 (3,7%) |  |
| Tremor | 2 (28,6%) | 1 (14,3%) | 0 | 0 | 3 (15%) | 0 | 0 | 0 |  | 5 (18,5%) | 1 (3,7%) | 0 | 0 |  |
| Myoclonus | 1 (14,3%) |  |  |  |  |  |  |  |  | 1 (3,7%) | 0 | 0 | 0 |  |
| Hypophosphataemia | 4 (57,1%) | 2 (28,6%) | 0 | 0 | 1 (5%) | 3 (15%) | 4 (20%) | 0 |  | 5 (18,5%) | 5 (18,5%) | 4 (14,8%) | 0 |  |
| Face edema | 0 | 0 | 0 | 0 | 0 | 1 (5%) | 0 | 0 |  | 0 | 1 (3,7%) | 0 | 0 |  |
| Depressed level of consciousness |  |  |  |  | 0 | 0 | 0 | 2 (10%) |  | 0 | 0 | 0 | 2 (7,4%) |  |

### Characteristics of manufactured CD19 CAR-T cells

Leukapheresis was performed in patients diagnosed with R/R B-cell malignancies who had blood T-cell counts above 150 cells/μL on the day of collection. For four patients (pt12, pt16, pt19, pt20), T-cells from haploidentical donors were used as the starting material for CAR-T cell manufacturing. The average volume of the apheresis product was 117 mL (range: 70–250 mL). On average, 5 × 10^9 (range: 0.015–64.6 × 10^9) PBMCs were isolated from the apheresis product. In seven cases, the PBMCs fraction was manually isolated and cryopreserved for later use; in the remaining cases, CAR-T cell production proceeded immediately. Activation and transduction of CD4□ and CD8□ lymphocytes were performed separately, followed by expansion in the presence of IL-7 and IL-15. The selected CD4□ and CD8□ T cells were transduced with a lentiviral vector BY19. The production process took between 7 to 31 days (median: 13 days). The resulting CAR-T cell products were administered immediately to 18 patients; for five patients, the products were cryopreserved until use. The quality of the initial material varied significantly, particularly regarding the content of naïve T cells (Tn) and stem cell memory T cells (Tscm), with median 28.3% (range 0% to 79%) for CD4□ T cells and median 25.8% (range 0% to 82%) for CD8□ T cells. Two patients (pt03, pt05) underwent immunomagnetic CD45RA selection of T cells prior to CAR-T manufacturing. Data on main characteristics of CAR-T products is shown in Figure 4. In vitro expansion over 12 days revealed a median fold increase of 13 (range: 0.24–188) for CD4□ cells and 11 (range: 0–195) for CD8□ cells.

**Figure 4.**
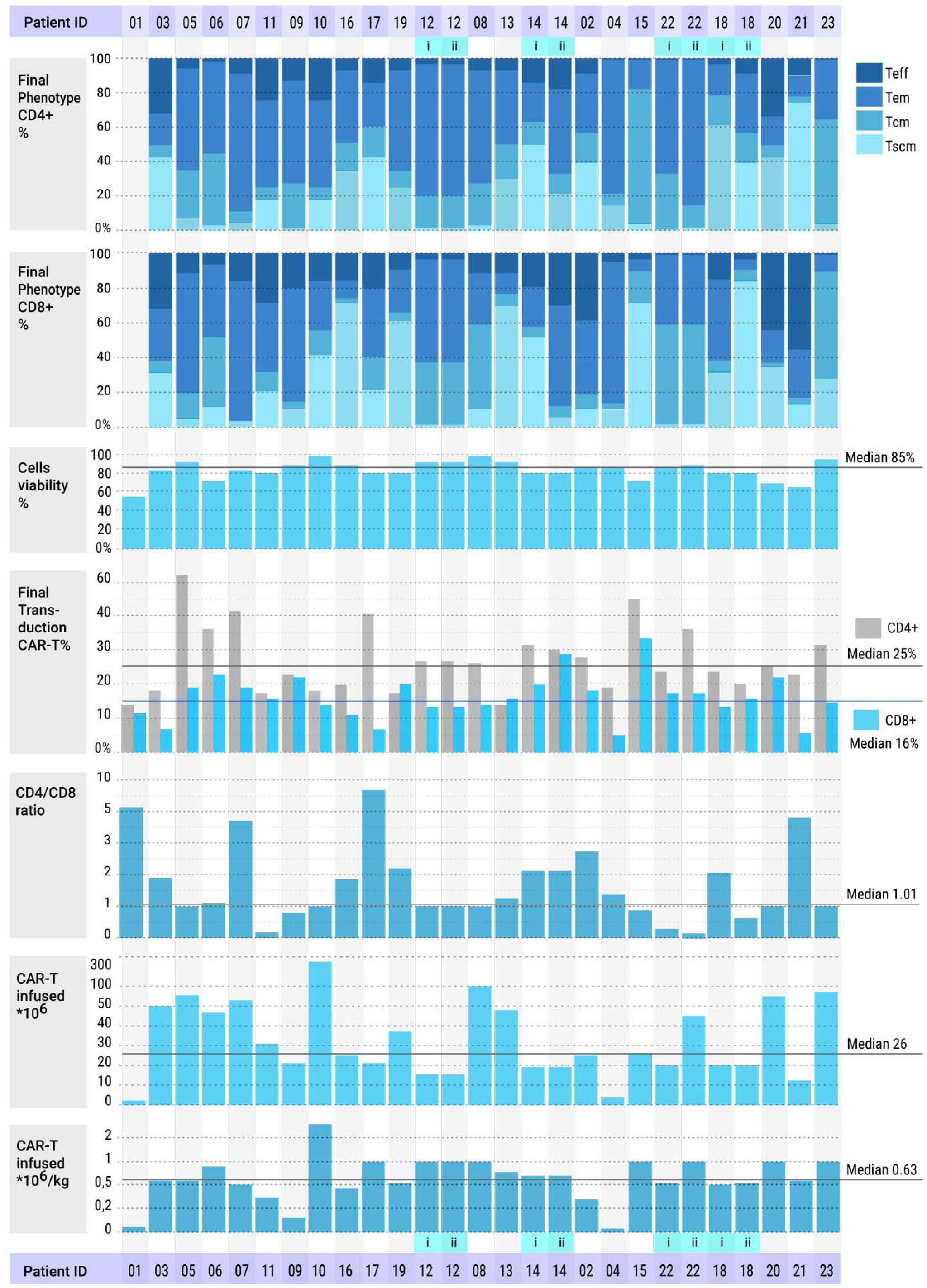
General characteristics and subpopulation analysis of the final CAR-T cell product. Teff, terminal effector T-cells; Tem, effector memory T-cells; Tcm, central memory T-cells; Tscm, stem cell memory T-cells;

The median cell viability was 84.5% (range: 55–96%). Transduction efficiency was higher for CD4□ lymphocytes, with a median of 24.5% (range: 13–63%), compared to CD8□ lymphocytes, which had a median of 16.35% (range: 6–33%). The desired CD4:CD8 ratio in the final product was 1:1; however, due to an insufficient total dose and limited CD8□ cell numbers, the actual CD4:CD8 ratio varied from 0.1 to 6.9, with a median of 1.03. The median CAR-T cell dose infused was 0.6 × 10^6 cells/kg (range: 0.015–2.8 × 10^6 cells/kg). Overall, the final CAR-T cell products contained a substantial fraction of memory T cells - Tscm and Tcm subsets — with median proportions of 44.4% (range: 5.2–84%) for CD4□ cells and 37.8% (range: 2.8–90%) for CD8□ cells, confirming the retention of a memory phenotype in the manufactured products (see Figure 4).

### Safety

No acute infusion-related toxicity was observed within the first 2 hours after CAR-T cell infusion. Overall, safety evaluations included 27 treatment episodes, as four patients received a second infusion of CD19 CAR T-cells. In the full safety cohort (n=27), 92.6% of patients (n=25) experienced any treatment-emergent adverse events (see Table 1). Patients developed expected toxicities, CRS, ICANS, cytopenias, and infections. CRS occurred in 71% of patients (n=5) at VRCCC and 65% (n=13) at BRC POHI, resulting in an overall CRS incidence of 67% (n=18) across both centers, with most cases being mild. Severe CRS (≥ grade 3) was observed in 19% of patients (n=5). The median time from infusion to CRS onset was 6 days (range: 1–13 days), and the median duration of CRS was 5 days (range: 1–17 days). Tocilizumab was administered after 12 CAR-T infusions (52.17%), with a median dose of 1 dose per patient (range: 1–7 doses).

ICANS occurred in 42.8% of patients (n=3) at VRCCC and 45% (n=9) at BRC POHI, resulting in an overall ICANS incidence of 44% (n=12) across both centers. Severe ICANS (≥ grade 3) was observed in 18.5% of patients (n=5) (see Table 1). The median time from infusion to ICANS onset was 8.5 days (range: 4–17 days), with a median duration of 3 days (range: 1–13 days). The clinical presentation of neurotoxicity was variable, ranging from mild to severe encephalopathy, focal neurological deficits, and ataxia, to more severe manifestations such as generalized seizures. All neurological symptoms and signs resolved completely over days to weeks, except for one patient who developed severe, irreversible neurological deficits and subsequently died 8 days after CAR-T cell infusion. Four patients developed transient disseminated intravascular coagulation (DIC), which was successfully managed with fibrinogen concentrate, cryoprecipitate, and coagulation factor concentrates. Grade 4 HLH occurred in two patients at VRCCC, and one patient at BRC POHI died due to unresponsive HLH (see Table 1). No GVHD were observed in any patient treated with haploidentical donor CAR-T cells. Steroids were administered to 10 patients (43.4%), with a median cumulative dose of 286 mg of dexamethasone (range: 8–1145 mg). Two patients died from severe infections in the context of CRS: one (patient 23) due to septic shock caused by pan-resistant Klebsiella pneumoniae and Acinetobacter baumannii, and another (patient 20) due to septic shock caused by multi-resistant Pseudomonas aeruginosa. Additional infections recorded included one case of invasive pulmonary aspergillosis, two bacterial pneumonias, two COVID-19 infections, one central line-associated bloodstream infection (CLABSI), one skin infection, one CMV reactivation, and one sepsis due to Klebsiella pneumoniae, all of which were successfully managed (see Table 1).

Cytopenias are a well-recognized side effect following CD19 CAR-T cell therapy. Neutropenia was graded according to the EHA/EBMT consensus [27], while anemia and thrombocytopenia were graded using CTCAE version 4.03. In brief, the majority of patients experienced early immune effector cell-associated hematologic toxicity (ICAHT), occurring in 81.5% (n=22) of cases, predominantly of high-grade severity— grade 3 in 7.4% (n=2) and grade 4 in 74.1% (n=20). Late ICAHT was observed in 29.6% (n=8) of patients, with grade 2 toxicity diagnosed in 5 patients (18.5%) and grade 4 in 3 patients (11.1%). Anemia was diagnosed in 26 patients (96%), with severe anemia recorded in 11 patients—grade 3 in 10 patients (37%) and grade 4 in 1 patient (3.7%). Thrombocytopenia was noted in 25 patients (92.5%), mostly of grade 3 (33%, n=9) and grade 4 (44.4%, n=12). By month 3, no patients exhibited severe hematological toxicity. All non-hematological adverse events were low grade and managed effectively. No cases of second primary malignancies have been documented during the observation period.

### Efficacy

The cutoff date for the updated analysis was June 17, 2024, resulting in a median follow-up of 15.4 months. Six patients responded to therapy, yielding an ORR of 85.7% (6/7; 95% CI: 42.1–99.6%), with five CR, corresponding to a CR rate of 71.4% (5/7; 95% CI: 29.0–96.3%) at VRCCC. At BRC POHI, 10 patients achieved CR, resulting in an overall CR rate of 76.9% (10/13 evaluable patients; 95% CI: 46.2–95.0%). The combined ORR across both centers was 80.0% (16/20; 95% CI: 56.3–94.3%), with a CR rate of 75.0% (15/20; 95% CI: 50.9–91.3%) observed at the first disease assessment on day 28. Among six patients with non-Hodgkin lymphoma, five responded to therapy, resulting in an ORR of 83.3% (5/6; 95% CI: 35.9– 99.6%) and a CR rate of 83.3% (5/6; 95% CI: 35.9–99.6%).

In the cohort of 14 evaluable patients with B-ALL, response assessment on day 28 following the first infusion of CD19 CAR-T cells showed an ORR of 78.6% (11/14; 95% CI: 49.2–95.3%). CR was achieved in 71.4% of B-ALL patients (10/14; 95% CI: 41.9–91.6%), with MRD-negative remissions detected in 57.1% (8/14; 95% CI: 28.9–82.3%) as determined by high-resolution flow cytometry. A second infusion of CD19 CAR-T cells was administered to four patients, with two of them responding (Figure 5). Patients with lymphoma who achieved CR demonstrated very durable responses, with no relapses reported at the time of manuscript writing. Eight patients with B-ALL who responded to CD19 CAR-T cell therapy proceeded to alloSCT, of whom six remain in CR at the latest follow-up. One patient (Pt03), who underwent lymphodepletion with decitabine, fludarabine, and cyclophosphamide and did not proceed to alloSCT, remains in CR without any further therapy. Median PFS was 23 months, and the median OS was not reached. 12 months PFS was 83.3% for patients with non-hodgkin lymphoma and 48.3% for patients with B-ALL (Figure 5).

**Figure 5.**
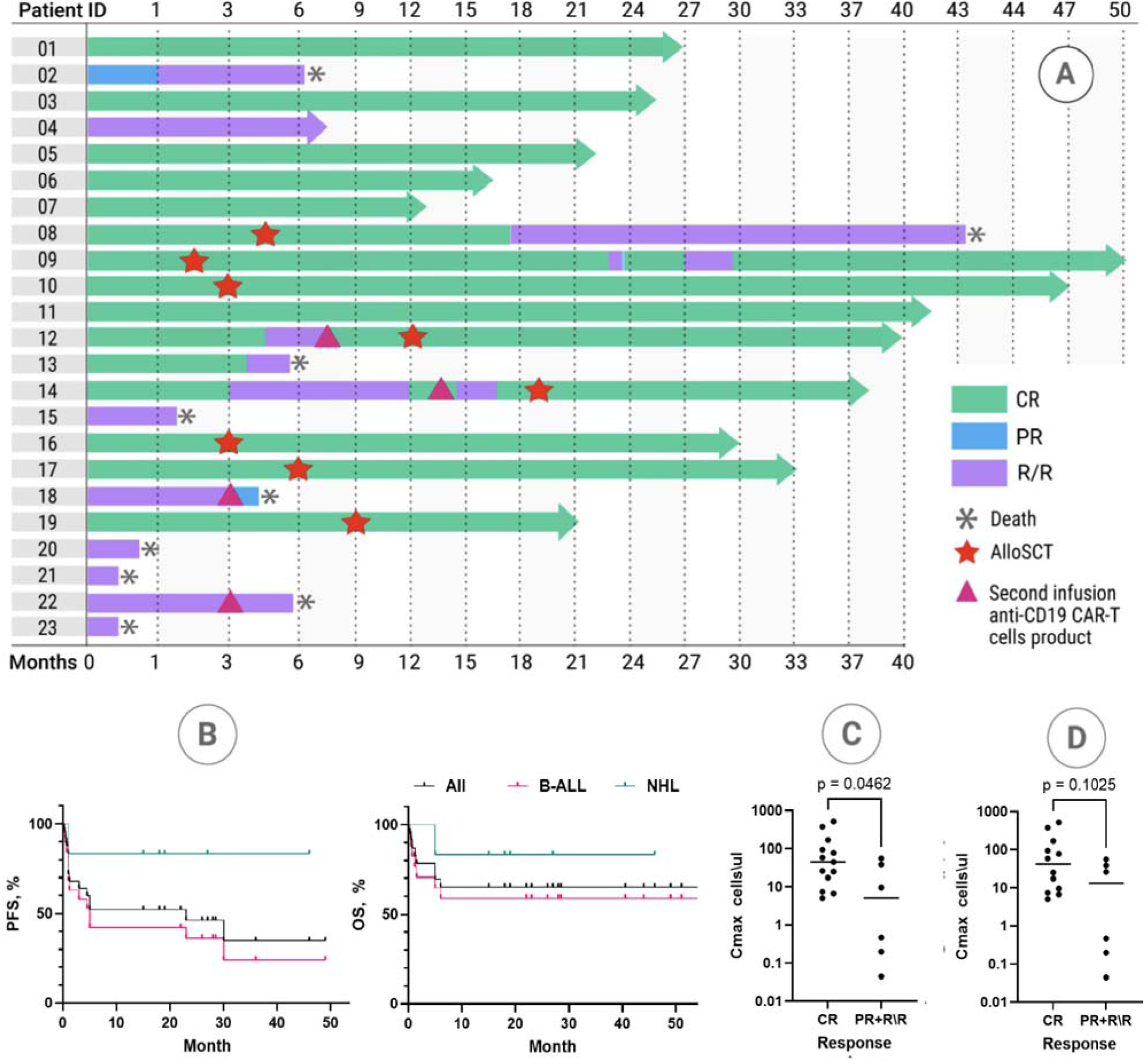
Clinical outcomes after CD19 CAR-T cell therapy. A) Swimmer’s plot illustrating responses and clinical courses of patients who underwent CD19 CAR T-cell therapy. B) Progression-free survival (PFS) and overall survival (OS) stratified by disease type. CR, complete response; PR, partial response; R/R, refractory/relapsed disease; B-ALL, B-cell acute lymphoblastic leukemia; NHL, non-Hodgkin lymphoma. C) Correlation between response at 1 month and CD19 CAR-T cell C_max_. The probability of achieving a response at 1-month post-infusion is positively correlated with C_max_ of CD19 CAR-T cells, indicating that higher CAR-T cell expansion is associated with improved early response rates (CR vs PR or R/R; p=0.0462). D) Correlation between 1-year PFS and CD19 CAR-T cell C_max_ (p=0.1330).

### Cellular kinetics of anti-CD19 CAR–T cells

CAR T-cell persistence was assessed in peripheral blood by flow cytometry at two or more time points after infusion in the majority of patients. At peak expansion, CAR T-cells comprised up to 48% of all viable nucleated cells (median: 11.6%; range: 1.5%–48.3%). Regardless of the CD4:CD8 ratio in the final CAR T-cell product, most patients exhibited predominant expansion of CD8 CAR T-cells after infusion. All evaluable patients, except two (Pt15 and Pt21), demonstrated CD19 CAR T-cell expansion in peripheral blood, with a median C_max_ of 21.7 cells/_μ_L (range: 0.045–1858 cells/_μ_L) and a median T_max_ of 10 days (range: 7–17 days) (Figure 6). A significant correlation was observed between CAR T-cells C_max_ and CR attainment (p = 0.0462), whereas no correlation was found with 1-year PFS (p = 0.1330) (Figure 5).

**Figure 6.**
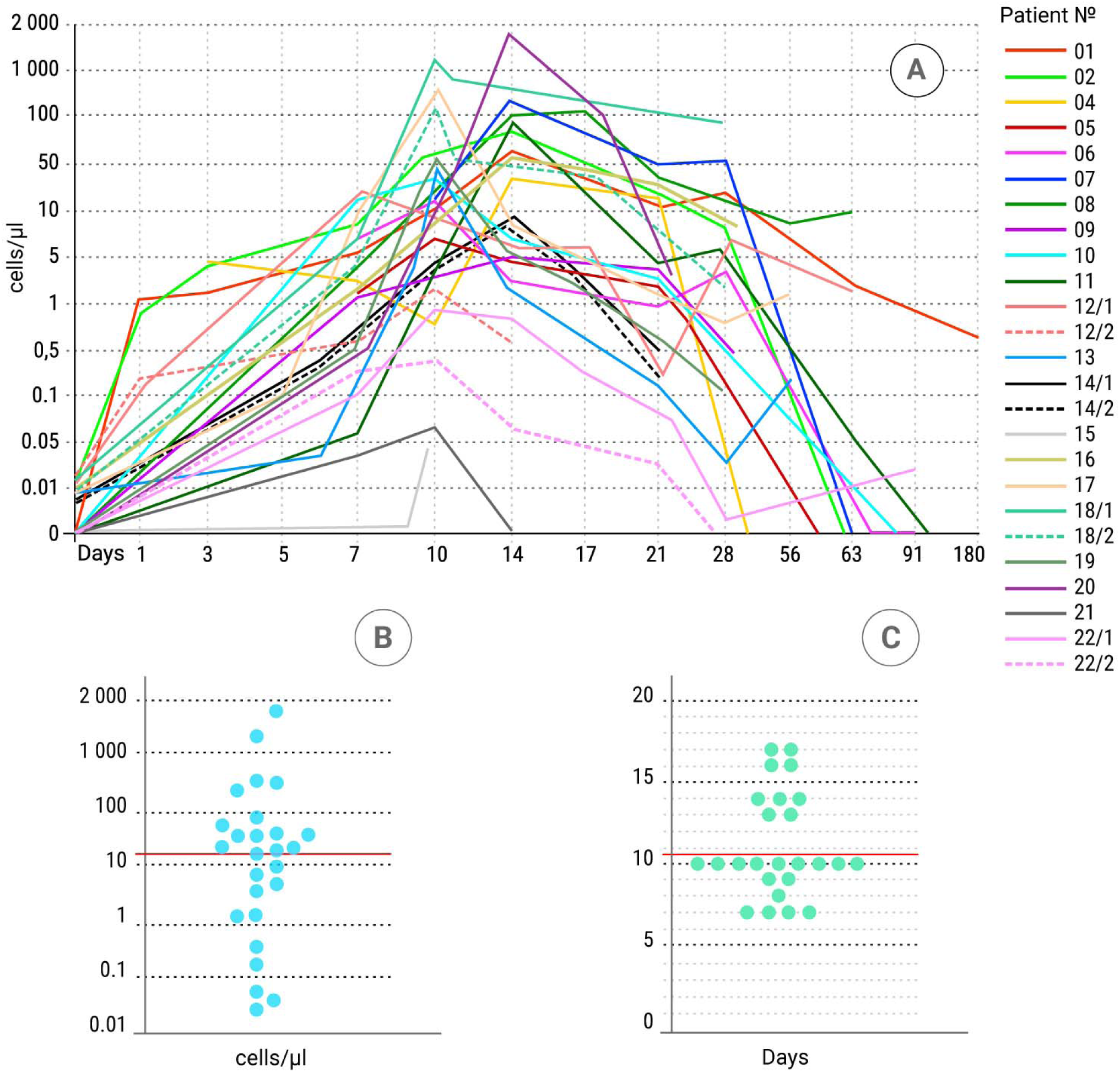
Pharmacokinetics of CD19 CAR-T cells in the peripheral blood of patients. A) Expansion and persistence of CD19 CAR-T cells over time. B) Peak concentration (C_max_) of CD19 CAR-T cells measured in peripheral blood, expressed as cells/µL. C) Time to maximum concentration (T_max_) of CD19 CAR-T cells in peripheral blood, expressed in days.

## DISCUSSION

This first-in-region phase I study demonstrates the feasibility, safety, and preliminary efficacy of an in-house manufactured CD19 CAR T-cell product (BY19) for patients with relapsed/refractory (R/R) B-cell malignancies in Belarus. Our experience provides a clinically actionable framework for developing academic CAR T-cell programs in low- and middle-income countries, where access to commercial cell therapies remains limited due to prohibitive costs, centralized logistics, and regulatory hurdles.

The BY19 product features a second-generation CAR construct with an IgG4 hinge, CD28 costimulatory domain, and CD3ζ was jointly developed by the Institute of Bioorganic Chemistry of NASB and BRC POHI and produced in an academic facility under GMP-adapted procedures at BRC POHI. The entire production cycle, including leukapheresis, transduction, expansion, and quality control, was completed in a median of 13 days. Both fresh and cryopreserved products were successfully transported from the manufacturing facility at BRC POHI to the clinical site at VRCCC, confirming the logistical and functional viability of academic manufacturing and inter-site transport within a national network.

Conducting scientific research and clinical testing of our CD19 CAR-T cell product occurred during a period when extensive data from various Phase I/II clinical trials exploring different anti-CD19 CAR designs were available. Among 39 studies that tested at least two dose levels of CAR-T cells, an association between dose administered and ORR or CR was observed in only 13 (33%) studies, raising questions about a strict dose-response relationship [28]. Notably, in these studies, the starting dose was comparatively lower (<30 million cells), whereas in studies showing no correlation between dose and disease response, the starting dose or dose level 1 (DL1) exceeded 50 million cells. Since the minimum effective dose often occurs well before the maximum tolerated dose in traditional phase I trials for hematological malignancies, there is a strong rationale to identify an optimal dose window—one that maximizes efficacy without increasing the risk of toxicity and adverse events [29]. Additionally, managing severe CRS and ICANS may lead to higher cumulative corticosteroid doses, which have been associated with significantly shorter PFS and OS [30]. Considering that most CD19 CAR-T cells studies achieved optimal clinical efficacy (greater than 70% ORR) at doses between 50 and 100 million cells, we selected these doses for our CD19 CAR T-cells study in adult patients [28].

Moreover, we incorporated decitabine (DAC) into the lymphodepletion regimen for adult patients prior to CAR T-cell infusion. DAC is a nucleoside analog whose phosphate group covalently binds to DNA methyltransferase (DNMT), inhibiting its activity. As a result, DAC functions as a hypomethylating agent (HMA) by promoting DNA demethylation [31]. Preclinical evidence demonstrates that preincubation of CD19+ lymphoma cell lines with DAC significantly enhances the cytotoxic capacity of anti-CD19 CAR T-cells [32]. One direct mechanism involves DAC-mediated upregulation of CD19 expression on lymphoma cells, while indirect in vivo effects may include disruption of the immunosuppressive tumor microenvironment [33–35]. In a small study, incorporating decitabine into lymphodepletion significantly improved response durability, resulting in longer PFS and OS in patients with B-ALL receiving CD19/CD22 CAR T-cells [36].

An additional advantage of DAC is its ability to penetrate the central nervous system, which could be highly valuable for improving outcomes in CNS lymphoma treated with CAR T-cell therapy, given its immunomodulatory activity discussed earlier [37]. The successful outcome of our PCNSL patient, who received decitabine-based lymphodepletion followed by anti-CD19 CAR T-cell therapy, supports further investigation into this approach. The toxicities associated with decitabine-based lymphodepletion combined with CAR T-cell therapy were manageable, with only two cases of CRS among seven patients and three cases of ICANS.

Interestingly, our CD19 CAR T-cells combined with DAC-based lymphodepletion demonstrated high efficacy against multiple types of non-Hodgkin lymphoma, comparing favorably to the response rates observed with FDA-approved products such as axicabtagene ciloleucel, lisocabtagene maraleucel, and tisagenlecleucel [38, 39]. The remission rates, EFS, and OS for patients with B-ALL in our study align with the lower-end results reported for other CD19 CAR T-cell products [13, 17, 39]. These outcomes may be attributed to a heavily pretreated patient population, including prior therapies with blinatumomab, high tumor burden, extramedullary disease such as CNS involvement, and colonization with multidrug-resistant gram-negative bacteria, which contributed to two sepsis-related deaths (Figure 2). This underscores that infections remain a significant challenge in highly pretreated hematological patients undergoing CAR T-cell therapy and could contribute to excessive mortality rate [40]. Our observations suggest that patients with higher C_max_ tend to achieve higher response rates. However, this study has limitations, including the small sample size and heterogeneity of the patient population. Consequently, we cannot draw definitive conclusions regarding safety and efficacy or establish conclusive correlative associations. Nonetheless, DAC-based lymphodepletion appears promising and warrants further investigation. To address these issues more robustly, ongoing recruitment across different disease cohorts is essential to gather sufficient data for definitive assessments of safety and efficacy across various types of lymphoid neoplasms.

## CONCLUSION

This phase I study establishes the feasibility and clinical potential of decentralized, academic CAR T-cell therapy in a resource-limited setting. The BY19 product, manufactured locally and infused at national cancer centers in Belarus, produced encouraging clinical responses with a manageable safety profile. The integration of decitabine into lymphodepletion may enhance antitumor activity and deserves further exploration, particularly in CNS lymphoma.

Our experience demonstrates that safe, effective, and scalable CAR T-cell therapy can be developed and delivered outside of high-income academic hubs. This model offers a compelling blueprint for improving access to transformative cell therapies globally and underscores the importance of continued investment in locally adaptable immunotherapy platforms.

## Data Availability

All data produced in the present study are available upon reasonable request to the authors

## AUTHOR CONTRIBUTIONS

Conceptualization, A.M. (Alexander Meleshko), A.M. (Alexandr Migas), D.D., M.K.; Investigation, All Authors; Writing – Original Draft, M.K.; Writing – Review & Editing, A.M. (Alexander Meleshko), A.M. (Alexandr Migas), D.D.; Funding Acquisition, A.M. (Alexander Meleshko), D.D. All authors have read and agreed to the published version of the manuscript.

## ACKNOWLEDGMENT

The work was supported by the State Committee on Science and Technology of Belarus (Agreement No. 201739). We sincerely appreciate the contributions of Professor Olga Aleinikova and Professor Natalya Konoplya in advancing CAR T-cell therapy from the preclinical stage to clinical investigation, as well as for their mentorship. We also extend our gratitude to the physicians, nurses, and staff at the BRC POHI and VRCCC laboratories for their dedicated routine work.

## DATA AVAILABILITY

The main data supporting the results in this study are available within the paper and the supplement information. All data generated in this study can be obtained from the corresponding authors upon reasonable request.

