## Supplementary figure 1 for "CD19 CAR T-cell (BY19) therapy for Pediatric and Adult Patients with Relapsed or Refractory B-Cell Neoplasms in Belarus: Phase 1 trial"

### Supplementary Information

**Supplementary Figure 1 - CAR mediated activation of NFkB (A,B,C) and NFAT (D,E,F) reporter genes.** EGFP expression kinetics of stimulated BY19 and 28Z CAR reporter cells - K562 (A and D), Raji (B and E) and CII (C and F). Data represent MFI ± SD. MOCK - untransduced cells.

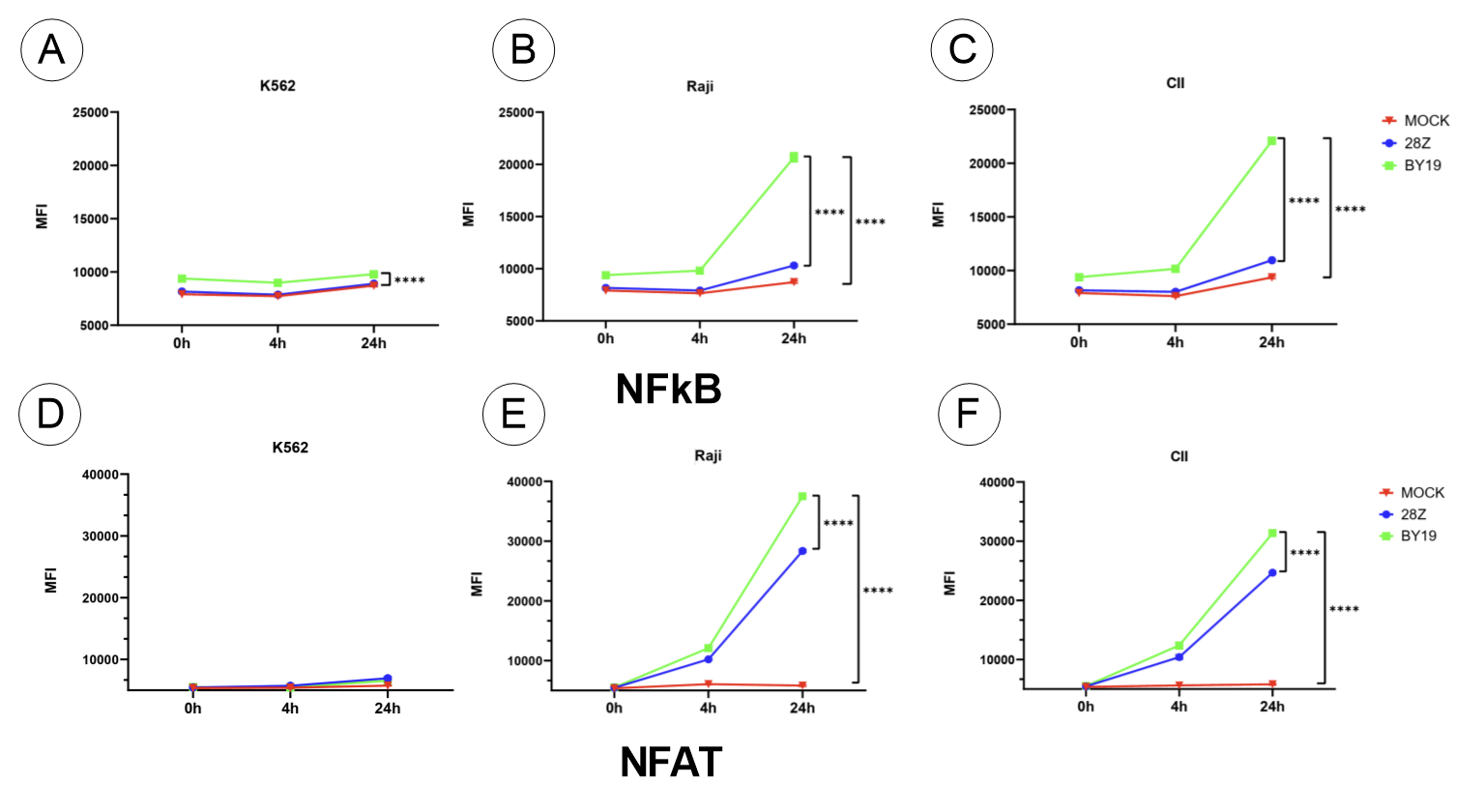
